# Pediatric HIV Hotspots in Kenya: Machine Learning and Geostatistical Analysis for Enhanced Case Finding

**DOI:** 10.64898/2026.04.24.26351710

**Authors:** Amobi Andrew Onovo, Boniface Makone, Fredrick Miruka, Edmon Obat, Peter Yegon

## Abstract

**Background:** Although Kenya’s HIV programme has long prioritized high-burden counties for intensified paediatric interventions, a critical evidence gap remains in developing integrated analytic frameworks that can objectively predict and validate paediatric HIV burden using data-driven models. We therefore developed and tested a framework that combines machine-learning (ML) prediction with geostatistical hotspot analysis, where a hotspot denotes a statistically significant spatial cluster of elevated paediatric HIV cases to strengthen data-driven surveillance and resource targeting.

**Methods:** National HIV testing data for children aged 0–14 years were analysed together with indicators from the 2022 Kenya Demographic and Health Survey. Multiple supervised ML algorithms were trained to predict the number of children living with HIV (CLHIV) across Kenya’s 47 counties. Model performance was evaluated using root-mean-square and mean-absolute error. The tuned Lasso-regression model demonstrated the best predictive accuracy and generated county-level estimates for October 2022 to June 2023. These predictions were subsequently assessed for spatial autocorrelation (Moran’s I) and validated using Getis-Ord Gi* statistics.

**Findings:** The model predicted 3160 newly identified CLHIV during the study period, compared with 3092 cases reported nationally. To account for differences in county population size, paediatric HIV incidence was calculated as cases per 10,000 children aged 0–14 years using 2023 census projections as the denominator. Incidence-based choropleth maps revealed that the highest reported burden was concentrated in Isiolo (11·2 per 10,000) and western Kenya (Homa Bay 7·7, Kisumu 3·6, Siaya 3·5), while model predictions identified additional high-incidence counties in eastern and northern regions. Significant spatial clustering was confirmed for both reported (z = 3·23, Moran’s I = 0·22, p = 0·001) and predicted (z = 4·92, Moran’s I = 0·37, p < 0·001) distributions. Thirteen counties, predominantly in western Kenya, were identified as statistically significant hotspots.

**Interpretation:** This study presents a validated methodological framework integrating ML prediction with geostatistical analysis for paediatric HIV surveillance. By expressing model outputs as population-adjusted incidence, the framework enables equitable comparison of paediatric HIV burden across counties of differing size, strengthening the evidence base for geographic prioritization and resource allocation.

**Funding:** This research received no specific grant from any funding agency in the public, commercial, or not-for-profit sectors.

## Introduction

Children remain disproportionately affected by inequities in HIV testing and treatment coverage, particularly across sub-Saharan Africa.^1,3^ Despite global ART scale-up and a steady decline in AIDS-related mortality, paediatric HIV continues to pose a major public-health challenge because of delayed diagnosis, limited paediatric formulations, and weak linkage to care.³ In 2022, an estimated 1·5 million children (aged 0–14 years) were living with HIV worldwide, nearly 89 % of them in sub-Saharan Africa.□ Only 57 % of these children received antiretroviral therapy (ART), compared with 77 % of adults, underscoring persistent disparities in the HIV care cascade.^1^,□

Kenya mirrors these global gaps. Despite strong national leadership and sustained PEPFAR support, substantial challenges remain in paediatric case identification and treatment initiation.□,^1^□ Although adult prevalence has declined, paediatric infections persist in pockets with high maternal HIV burden and historical prevention-of-mother-to-child-transmission (PMTCT) gaps.□,^1^□ National programmes have already prioritised high-burden counties for intensified action, yet such targeting often depends on retrospective, aggregated data. There remains a need for predictive, objective, and reproducible analytic frameworks that can anticipate paediatric HIV burden and validate spatial clusters of infection in near real-time.

Geographic targeting is an established strategy for optimizing limited health-system resources.□ Mapping spatial heterogeneity in HIV prevalence allows policymakers to direct testing and treatment toward the most affected communities, improving health equity and impact.□,□ Spatial epidemiology also exposes contextual drivers, such as poverty, migration, and service access that are often masked in aggregate reporting.^1^□ Building on these programme efforts requires advanced analytic methods that can dynamically model and validate geographic risk patterns to guide precision interventions.

Machine-learning approaches provide this capability by revealing complex, non-linear relationships between demographic, behavioural, and health-system factors.□,^1^□ When integrated with geostatistical analysis, ML enables both prediction of case counts and validation of *hotspots*, defined here as statistically significant spatial clusters of elevated case density identified through Getis-Ord Gi* statistics.D This integration advances surveillance from descriptive mapping toward predictive epidemiology, allowing model outputs to verify and prioritize regions for intensified paediatric interventions.

### Study aims

This study sought to develop and validate a methodological framework that combines supervised machine-learning prediction with spatial hotspot analysis to enhance paediatric HIV surveillance in Kenya. The specific objectives were to identify the machine-learning model with the best predictive performance for estimating county-level paediatric HIV cases, to validate spatial clusters using Moran’s I and Getis-Ord Gi* statistics, and to demonstrate how this integrated framework can support ongoing surveillance, strategic planning, and resource optimization.

By linking national testing data with population- and system-level indicators, the framework offers a replicable, scalable approach for data-driven epidemic intelligence. It illustrates how combining ML, and spatial statistics can enhance surveillance platforms such as Kenya’s HIV Estimates Model,^1^□ transforming predictive analytics into actionable epidemiologic insights for paediatric HIV control.

## Methods

### Study Design and Setting

A dual analytical approach integrating machine-learning (ML) and geostatistical techniques was used to identify paediatric HIV hotspots across Kenya’s 47 counties. The analysis was conducted within a cross-sectional framework using county-level aggregate data from national HIV testing records and the 2022 Kenya Demographic and Health Survey (KDHS). Kenya, located in East Africa, has an estimated population density of 92·3 people per km^2^ in 2023 and approximately 20 million children under 15 years.□,□ the country’s devolved governance system provides a useful structure for sub-national surveillance. According to the 2018 Kenya HIV Estimates Report, Homa Bay, Siaya, Kisumu, Migori, and Nairobi counties have historically reported the highest HIV burden.^1^□

### HIV Testing Delivery Context

Kenya’s national HIV Testing Services (HTS) programme, supported by PEPFAR, the Global Fund, and other partners, implements facility and community-based testing strategies to identify people living with HIV and link them to care. Testing data are routinely disaggregated by age, sex, test result, and testing modality, captured through client registers, electronic medical records, and the DHIS 2 system. The country has adopted a three-test rapid algorithm in line with WHO’s 2019 guidance to improve diagnostic accuracy, replacing the previous two-test strategy.^11^ Testing and linkage activities are conducted by trained facility-based providers and community counsellors.

### Data Sources and Target Population

The study combined HTS programme data from Oct 1, 2022, to June 30, 2023, with county-level indicators from the 2022 KDHS (figure 2). The target population comprised children aged 0–14 years residing in Kenya who were tested for HIV during the study period, as captured in national aggregate HTS datasets. Analyses focused on newly diagnosed paediatric HIV cases to examine geographic disparities in detection and guide targeted interventions.

The outcome variable was the number of newly identified children living with HIV (CLHIV) per county. Eighteen explanatory variables were included, spanning maternal and child health, PMTCT seropositivity, nutrition, antenatal care attendance, skilled birth attendance, vaccination, malaria prophylaxis, sexual behaviour, gender-based violence, and socio-economic indicators. All KDHS and programme variables were aggregated at the county level; no individual-level data were accessed or analysed. This ensured comparability across regions and alignment with Kenya’s sub-national surveillance framework.

### Geo-Data

Spatial polygons were obtained from DIVA-GIS and the Humanitarian Data Exchange (HDX), both licensed under Creative Commons 4·0. All projections used the World Geodetic System 1984 (WGS 84) coordinate reference system. Analyses and map production were conducted in ArcGIS Pro v3·1 (ESRI, CA, USA).

### Data Pre-processing and Missing-Data Treatment

Data were cleaned and harmonized before modelling. Missing values were imputed using Predictive Mean Matching (PMM), which replaces missing entries with observed values from cases with similar predicted scores. This method preserves distributional properties and relationships among variables, improving model reliability. Variables with a variance-inflation factor (VIF) > 5 were excluded to reduce multicollinearity.

### Descriptive and Distributional Analysis

Prior to model development, univariate descriptive statistics were computed for the outcome variable and all 19 candidate predictors across the 47 Kenyan counties. For each variable, we reported the arithmetic mean with standard deviation, median with interquartile range, minimum–maximum range, and skewness using the adjusted Fisher–Pearson standardised moment coefficient (Table 1). Skewness was categorised following conventional thresholds as approximately symmetric (|skew| < 0·5), moderate (0·5–1·0), or high (> 1·0). To complement the numerical summaries, the empirical univariate distribution of each variable was visualised using Gaussian kernel density estimation with Scott’s rule bandwidth selection (Figure 2). The kernel density plots allowed direct visual assessment of distributional shape, modality, and the position of the mean relative to the median as an indicator of skewness.

**Table 1.** Descriptive statistics of the outcome variable and candidate predictors across 47 Kenyan counties.

| N o. | Variable | Domain | N | Mean (SD) | Median [IQR] | Range | Skew | Interpretation |
| --- | --- | --- | --- | --- | --- | --- | --- | --- |
| 1 | **Paediatric HIV seropositivity (%)** | **Outcome** | 47 | **2.05 (1.20)** | **2.14 [1.17]** | **0.0–4.9** | **-0.08** | **Symmetric** |
| 2 | PMTCT HIV seropositivity (%) | HIV burden | 47 | 1.60 (1.54) | 1.38 [0.72] | 0.0–6.8 | +2.25 | High |
| 3 | ANC from skilled provider (%) | Maternal health | 47 | 97.07 (5.08) | 98.80 [1.90] | 76.4–100.0 | -3.18 | High |
| 4 | Four or more ANC visits (%) | Maternal health | 47 | 63.28 (11.66) | 65.10 [13.30] | 31.2–82.2 | -0.76 | Moderate |
| 5 | Iron supplementation in pregnancy (%) | Maternal health | 47 | 88.84 (8.80) | 91.20 [6.35] | 48.0–98.2 | -3.02 | High |
| 6 | Neonatal tetanus protection (%) | Maternal health | 47 | 74.21 (10.27) | 75.10 [17.85] | 53.6–91.5 | -0.09 | Symmetric |
| 7 | Skilled birth attendance (%) | Maternal health | 47 | 86.74 (13.44) | 92.50 [10.45] | 52.6–99.4 | -1.45 | High |
| 8 | Facility-based delivery (%) | Maternal health | 47 | 79.48 (15.13) | 84.40 [17.35] | 43.2–99.1 | -0.99 | Moderate |
| 9 | Postnatal check within 2 days (%) | Maternal health | 47 | 71.21 (14.88) | 75.00 [20.45] | 37.0–90.5 | -0.70 | Moderate |
| 10 | Severe stunting, children <5 (%) | Child health | 47 | 4.28 (2.42) | 3.80 [2.25] | 0.6–13.4 | +1.67 | High |
| 11 | Children <5 slept under ITN (%) | Child health | 47 | 51.94 (23.84) | 57.60 [40.90] | 8.6–83.8 | -0.55 | Moderate |
| 12 | Three or more SP/Fansidar doses (%) | Child health | 47 | 12.95 (15.56) | 4.90 [23.60] | 0.0–59.1 | +1.19 | High |
| 13 | Two or more partners, past 12 mo (%) | Sexual behaviour | 47 | 2.89 (2.04) | 2.70 [2.40] | 0.0–11.0 | +1.50 | High |
| 14 | Non-cohabiting sex, past 12 mo (%) | Sexual behaviour | 47 | 16.11 (6.80) | 18.00 [7.90] | 0.5–27.9 | -0.65 | Moderate |
| 15 | Men never tested for HIV (%) | HIV testing | 47 | 30.90 (10.92) | 29.80 [15.05] | 10.8–60.5 | +0.58 | Moderate |
| 16 | Women never tested for HIV (%) | HIV testing | 47 | 17.83 (13.54) | 14.00 [9.75] | 4.9–79.0 | +2.72 | High |
| 17 | Women aged 15–49 owning a house (%) | Socioeconomic | 47 | 64.79 (14.03) | 66.40 [17.60] | 38.9–92.2 | +0.03 | Symmetric |
| 18 | Physical violence since age 15 (%) | Gender-based violence | 47 | 33.15 (11.76) | 34.60 [15.80] | 8.6–62.2 | -0.03 | Symmetric |
| 19 | Physical violence, past 12 mo (%) | Gender-based violence | 47 | 12.14 (5.63) | 11.00 [6.55] | 2.6–24.8 | +0.64 | Moderate |
| 20 | Sexual violence, past 12 mo (%) | Gender-based violence | 47 | 5.82 (3.47) | 5.60 [4.15] | 0.4–16.6 | +0.88 | Moderate |

Data source: 2023 PEPFAR programme data and 2022 Kenya Demographic and Health Survey.
All variables are expressed as county-level percentages. The outcome variable (paediatric HIV seropositivity) represents the percentage of children aged 0–14 years who tested HIV-positive among those tested through PEPFAR-supported services in 2023. Mean (SD) = arithmetic mean with standard deviation; Median [IQR] = median with interquartile range; Range = minimum–maximum. Skewness was categorised as symmetric ( $|\text{skew}| < 0.5$ ), moderate ( $0.5–1.0$ ), or high ( $> 1.0$ ); high-skew values are shown in red. The outcome variable is highlighted in coral.
Abbreviations: ANC = antenatal care; IQR = interquartile range; ITN = insecticide-treated net; PMTCT = prevention of mother-to-child transmission; SD = standard deviation; SP = sulphadoxine–pyrimethamine.

Together, Table 1 and Figure 2 provided three analytic functions. First, they documented the central tendency, dispersion, and range of every modelled variable, establishing baseline characteristics of the Kenyan county-level dataset for reviewer and reader reference. Second, they identified variables with pronounced positive or negative skewness and ceiling effects, particularly among maternal health indicators with near-universal coverage (antenatal care from a skilled provider, iron supplementation, skilled birth attendance) and HIV testing variables with strong right-tails (women never tested for HIV, PMTCT seropositivity) that would violate the normality assumptions of ordinary least-squares regression. Third, the distributional characteristics directly informed the decision to adopt penalised regression approaches (Ridge, Lasso, and Elastic Net), which are robust to non-normality and multicollinearity, for the primary predictive modelling described below.

### Machine-Learning Workflow

Three penalized regression algorithms (Ridge, Lasso, and Elastic Net) were implemented to model paediatric HIV case counts. These regularized models were chosen for their suitability in high-dimensional data, their ability to address multicollinearity, and their interpretability for policy applications. Ridge regression adds an L2 penalty to stabilize estimates; Lasso applies an L1 penalty to perform variable selection; Elastic Net combines both to capture correlated predictors and encourage group selection.

All analyses were conducted in Python 3·11 with *scikit-learn 1·4*. Numerical features were standardized (mean = 0, SD = 1). The dataset was split 70: 30 into training and testing subsets. Hyperparameters (λ penalty and α mixing ratio) were tuned by grid search with ten-fold cross-validation. Model performance was evaluated using Root Mean Squared Error (RMSE) and Mean Absolute Error (MAE); the model with the lowest error metrics was selected for spatial validation.

### Model Validation and Sensitivity Analysis

Predictive reliability of the tuned Lasso model was assessed by comparing the county-level distributions of predicted and reported paediatric HIV case counts (n = 47 per group) using complementary frequentist and Bayesian approaches. The frequentist comparison used Welch’s two-sample t-test, with Hedges’ g as the standardised effect size. To assess the strength of evidence supporting the null hypothesis of no distributional difference, a Bayesian sensitivity analysis was performed using a Jeffreys–Zellner–Siow (JZS) Cauchy prior on the standardised effect size (scale r = 0·71), reporting the Bayes factor in favour of the null hypothesis (BF□□) and the 95% highest-density interval (HDI) of the posterior distribution of the mean difference. Results are presented in Figure 4.

To quantify county-level uncertainty around individual predictions, a residual-based bootstrap procedure was implemented. For each county, the empirical prediction residual was computed as the difference between the reported and predicted case count from the tuned Lasso model (reported minus predicted). In each of 2000 bootstrap iterations (seed = 20260420), a residual was drawn at random with replacement from the empirical residual distribution across all 47 counties and added to the fixed point prediction to generate one sample from that county’s predictive distribution. Ninety-five percent prediction intervals were computed as the 2·5th and 97·5th percentiles of the resulting per-county bootstrap distribution, with lower bounds truncated at zero given the non-negative nature of count outcomes. This procedure preserves the point predictions from the primary Lasso model while using the empirical distribution of observed prediction errors to quantify uncertainty. Per-county point estimates with 95% prediction intervals are reported in Supplementary Table 2 and visualised in Supplementary Figure 1, which presents, for each county, the Lasso point prediction, the 95% prediction interval, and the reported case count with a connector segment between the predicted and reported values to allow direct visual assessment of county-level discrepancies alongside uncertainty bounds.

### Spatial Analysis

Predicted and reported county-level CLHIV counts were georeferenced and analysed using three complementary approaches. First, county-level paediatric HIV incidence was calculated as the number of newly identified cases per 10,000 children aged 0–14 years, using 2023 Kenya National Bureau of Statistics population projections as the denominator. Incidence-based choropleth maps were then produced to illustrate the spatial distribution of paediatric HIV burden, thereby adjusting for inter-county differences in paediatric population size. Second, global spatial autocorrelation was assessed using Moran’s I statistics to quantify clustering, dispersion, or randomness in paediatric HIV distribution (–1 = perfect dispersion; 1 = perfect clustering). Third, hotspot analysis was conducted using the Getis-Ord Gi* statistics to identify counties with significantly high or low clustering of paediatric HIV cases. Counties with high z-scores and neighboring counties with similarly elevated values were classified as *hotspots*, whereas negative z-scores indicated *cold-spots*. Two side-by-side choropleth maps (reported versus predicted incidence) were produced at 300 dpi to facilitate visual comparison of spatial patterns.

### Ethics

The analysis was conducted using secondary data from Kenya’s national HIV testing programme and population-based surveys, including the 2022 Kenya Demographic and Health Survey. In accordance with Kenya’s national HIV programme policy, informed consent had been obtained from all clients at the time of HIV testing. Only aggregated county-level data were analysed in this study. No individual or personally identifiable information was accessed, and therefore ethical approval was not required for this secondary data analysis. The datasets used in this study are publicly available online: the PEPFAR data can be accessed through the PEPFAR Panorama Spotlight (https://data.pepfar.gov), and the 2022 Kenya DHS data are available from the DHS Program website (https://dhsprogram.com/).

## Results

The analytical workflow (figure 1) outlines the sequential steps of data integration, machine-learning model development, and geostatistical validation. Descriptive statistics for the outcome variable and all 19 candidate predictors across the 47 Kenyan counties are presented in Table 1. The univariate kernel density distributions of these variables are shown in Figure 2 and show considerable variability across counties. Several indicators exhibited pronounced positive skewness, including PMTCT HIV seropositivity (skew = +2·25), women never tested for HIV (+2·72), severe stunting in children under five (+1·67), and two-or-more sexual partners in the past 12 months (+1·50), while three maternal health indicators showed strong negative skewness reflecting ceiling effects at near-universal coverage: ANC from a skilled provider (−3·18), iron supplementation in pregnancy (−3·02), and skilled birth attendance (−1·45). Other indicators such as facility delivery and neonatal tetanus protection were approximately symmetric. These distributional characteristics, detailed in Table 1 and Figure 2, informed the data-preprocessing procedures and penalised regression model selection applied before training.

**Figure 1:**
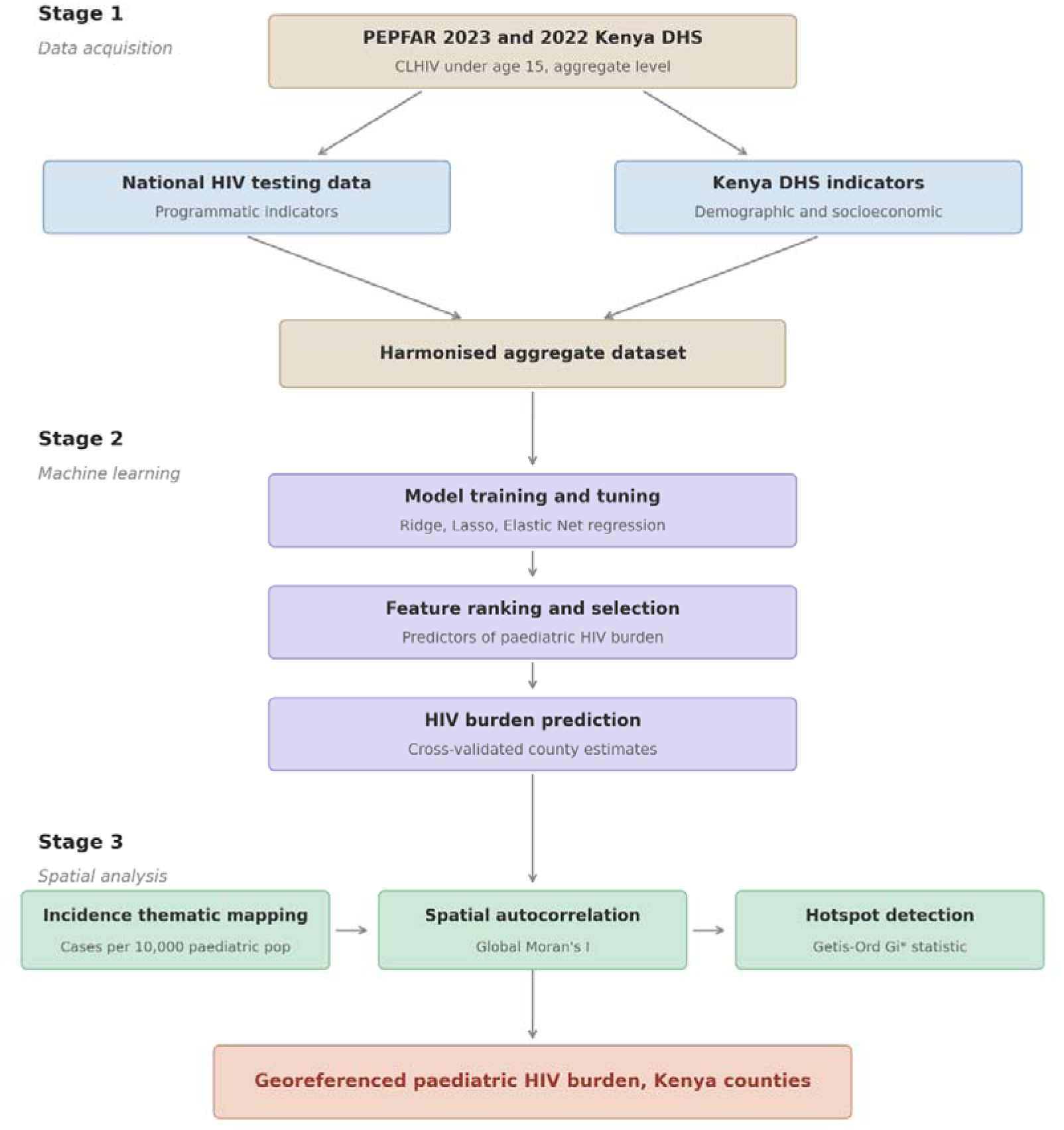
Analytical workflow for identifying paediatric HIV hotspots in Kenya. Data from Kenya’s national HIV testing programme and population-based surveys were integrated to create a unified analytical dataset. Machine-learning models were developed to estimate county-level paediatric HIV case counts across Kenya’s 47 counties. The predicted case counts were then incorporated into geostatistical analyses, including spatial autocorrelation and hotspot detection, to identify counties with significant paediatric HIV clustering. This integrated framework supports geographically targeted interventions and efficient resource allocation for epidemic control.

**Figure 2:**
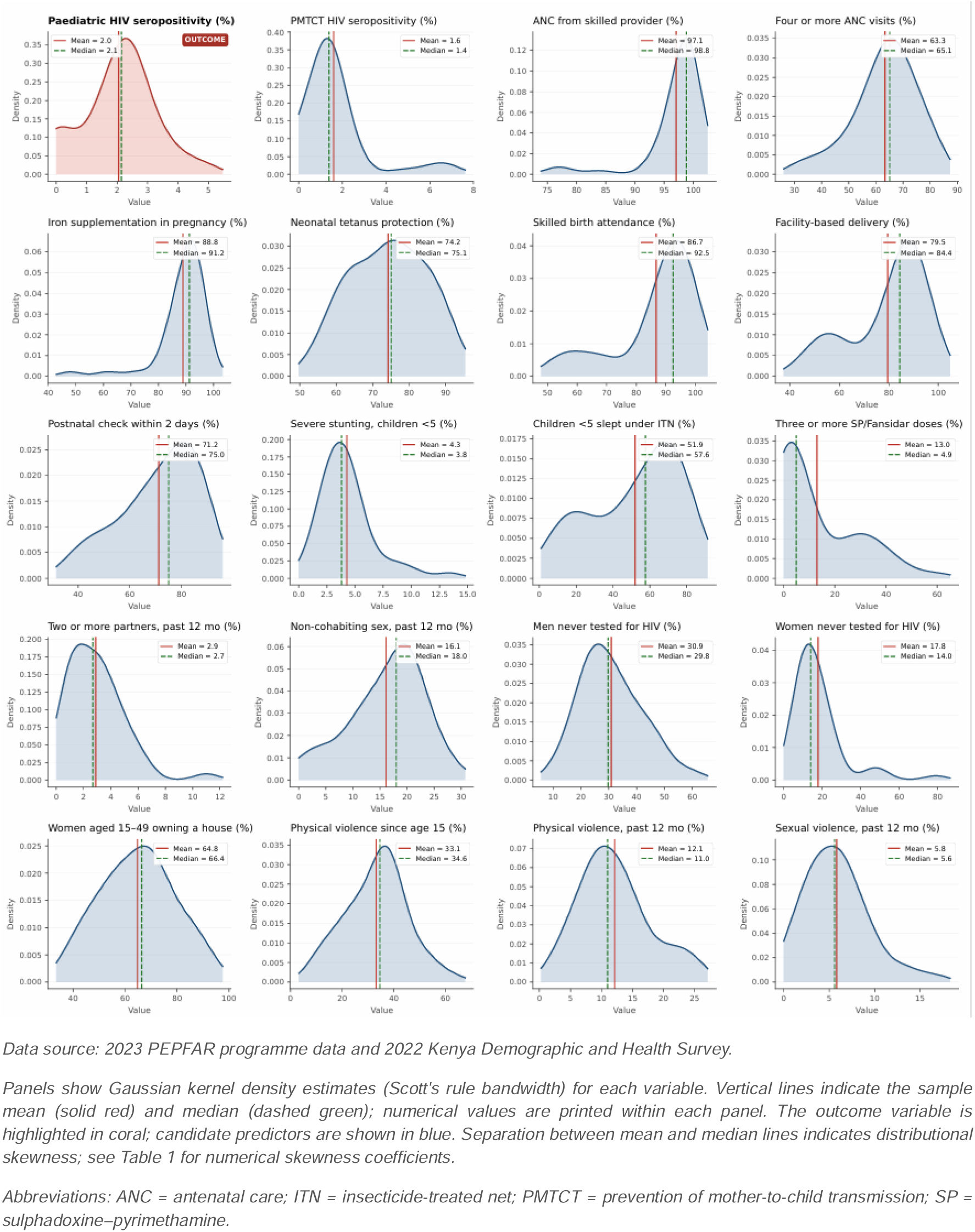
Kernel density distributions of the outcome variable and 19 candidate predictors across 47 Kenyan counties.

Across the supervised ML models tested (figure 3a), the tuned Lasso regression achieved the best overall predictive performance for estimating paediatric HIV cases, yielding the lowest root mean square error (RMSE = 0·122) and mean absolute error (MAE = 0·099). By contrast, Ridge and Elastic Net models showed higher error values. Feature-importance analysis highlighted PMTCT HIV seropositivity as the strongest positive predictor, followed by stunting, number of doses of sulphadoxine/pyrimethamine (Fansidar), and antenatal care (ANC) visits. Factors such as men never tested for HIV and recent experiences of sexual violence had strong negative coefficients, reflecting their association with poorer paediatric HIV outcomes. These findings emphasize the influence of maternal and household-level factors on paediatric HIV case detection and the potential value of strengthening family-centered testing strategies.

**Figure 3:**
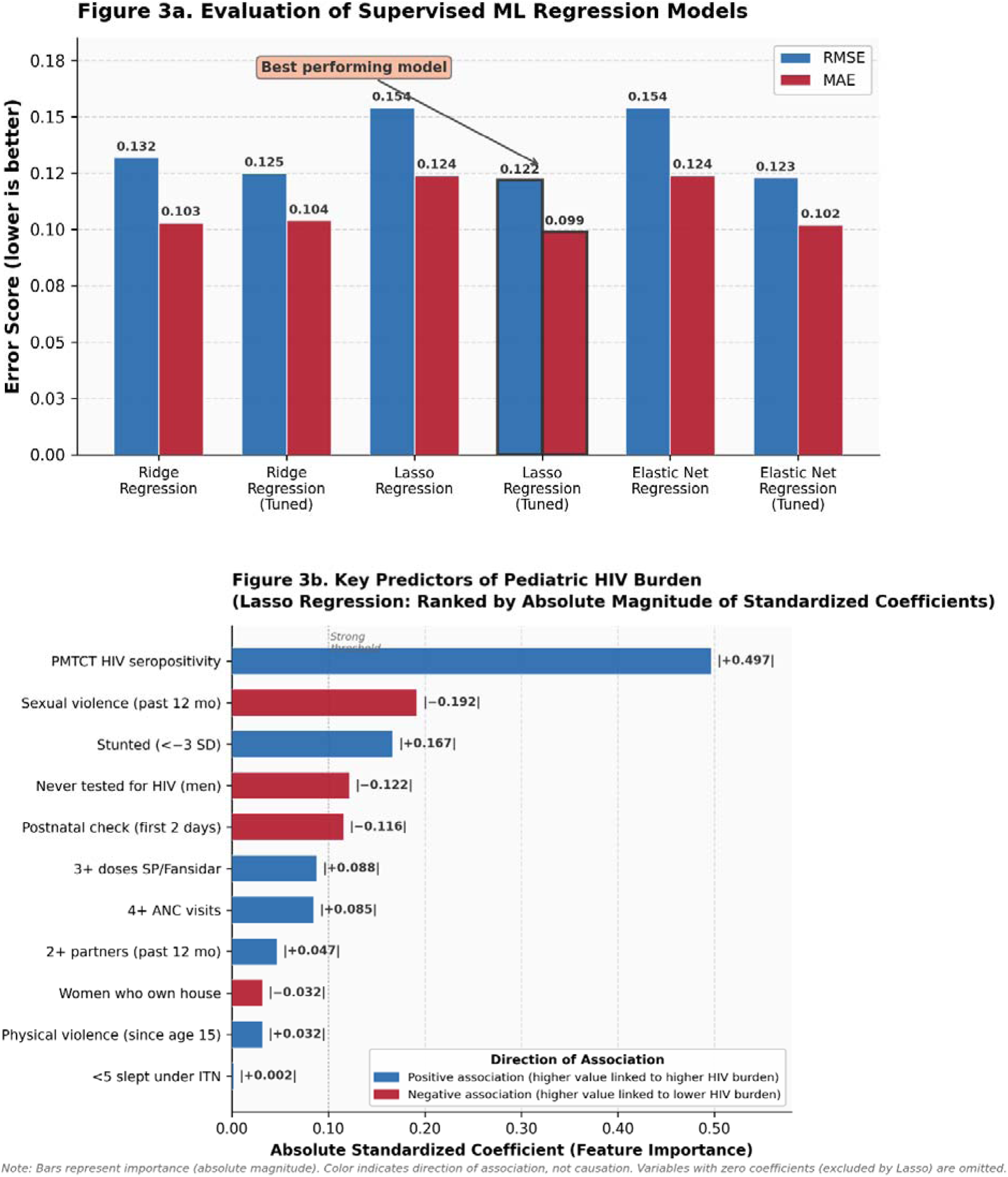
(a) Evaluation of supervised ML regression models for predicting paediatric HIV cases.

Model validation (figure 4) demonstrated high agreement between predicted and reported programme data. The tuned Lasso predictions (mean = 67·23, SD = 22·79) closely matched reported values (mean = 65·79, SD = 23·48). Welch’s two-sample t-test showed no significant difference between the distributions (t = 0·11, df = 86·99, p = 0·911), with a negligible effect size (Hedges’ g = 0·02; 95% CI −0·38 to 0·42). Bayesian sensitivity analysis using a JZS Cauchy prior (r = 0·71) yielded log(BF□□) = 1·52 (BF□□ ≈ 4·57), representing moderate evidence in favour of the null hypothesis of no difference between predicted and reported distributions; the 95% highest-density interval (HDI) of the posterior mean difference was −26·31 to 21·74 cases, confirming the model’s predictive reliability. Beyond the distributional comparison in Figure 4, county-level uncertainty was quantified through residual-based bootstrap prediction intervals (B = 2000; Supplementary Table 2 and Supplementary Figure 1). Across the 47 counties, the 95% prediction interval covered the reported case count in 45 counties (empirical coverage 95·7%, closely matching the nominal 95% level), indicating that bootstrap-derived uncertainty bounds were appropriately calibrated. The median prediction interval width was 159 cases. Two counties fell outside their 95% prediction intervals: Homa Bay, where the reported case count (399) exceeded the upper bound of the prediction interval (380), consistent with localised transmission intensity that exceeds what is predicted by county-level demographic and programmatic indicators alone; and Siaya, where the reported case count (148) fell below the lower bound (189), potentially reflecting under-detection or differential reporting coverage relative to underlying paediatric HIV burden. As shown in Supplementary Figure 1, these two counties sit at opposite edges of the reported–predicted distribution (Homa Bay above its upper bound, Siaya below its lower bound), and the directions of their outlying values carry distinct programmatic interpretations detailed in the Discussion. Both are among Kenya’s historically recognised high-burden paediatric HIV settings and both warrant continued prioritisation for intensified case-finding and targeted programming.

The ML model estimated 3160 newly identified paediatric HIV cases between October 2022 and June 2023, a modest increase from the 3092 cases reported by the national HTS programme. Figure 5 presents county-level paediatric HIV incidence (cases per 10 000 children aged 0–14 years) for both reported and predicted distributions. Among reported cases, Isiolo exhibited the highest incidence (11·2 per 10 000), followed by Homa Bay (7·7), Kisumu (3·6), and Siaya (3·5). The model-predicted incidence map revealed strong spatial concordance with reported patterns, with Isiolo (6·5), Siaya (5·7), and Homa Bay (5·3) ranked highest, while also identifying elevated predicted incidence in several counties where reported incidence was comparatively low, including Tana River (predicted 4·2 versus reported 1·0), Lamu (4·2 versus 2·8), and Vihiga (3·3 versus 1·2), suggesting potential under-detection in routine surveillance and opportunities for targeted testing expansion. By expressing case counts relative to the underlying paediatric population, the incidence-based maps distinguish counties with genuinely high per-capita burden from those with high absolute counts driven primarily by large population denominators.

**Figure 4:**
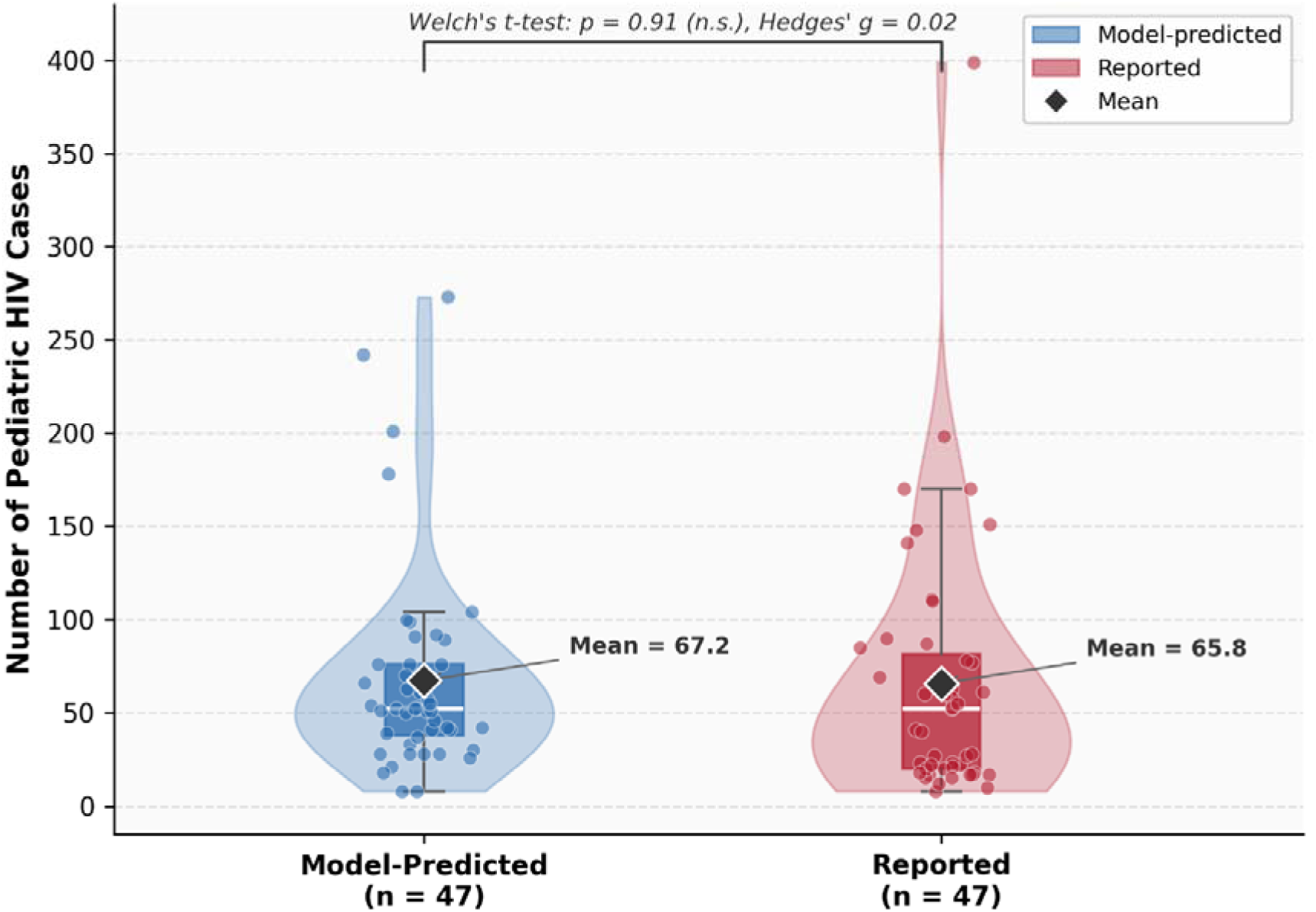
Comparative analysis of predicted and reported paediatric HIV cases across 47 Kenyan counties.

**Figure 5:**
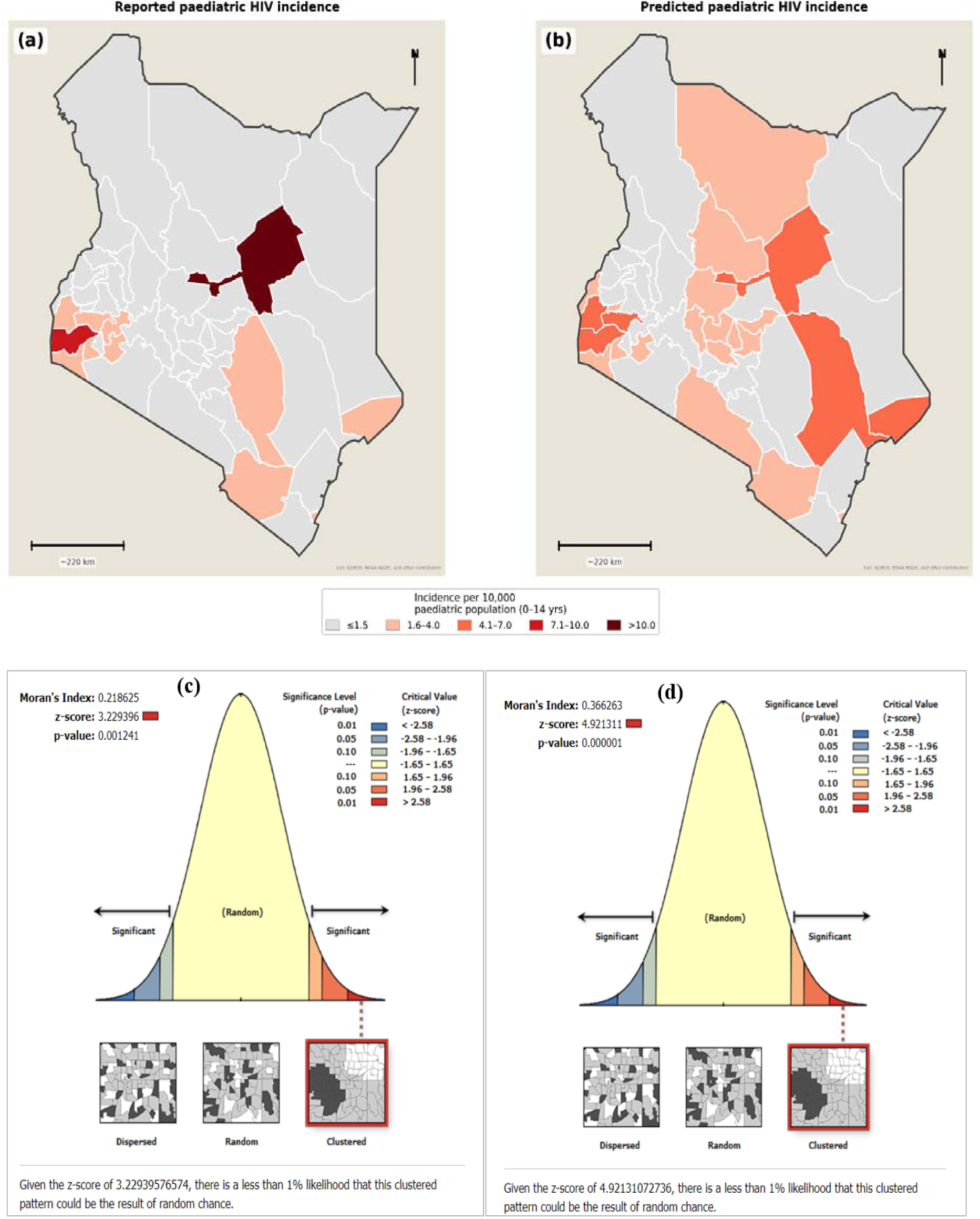
Geographic distribution and spatial autocorrelation of paediatric HIV across 47 Kenyan counties. (a) Reported paediatric HIV incidence (cases per 10,000 children aged 0–14 years) from the national HIV testing programme, October 2022 – June 2023. (b) Machine-learning predicted paediatric HIV incidence (cases per 10,000 children aged 0–14 years) as of end of June 2023. (c) Moran’s I scatter plot of reported paediatric HIV cases, showing global spatial autocorrelation. (d) Moran’s I scatter plot of predicted paediatric HIV cases, showing global spatial autocorrelation.

**Figure 6:**
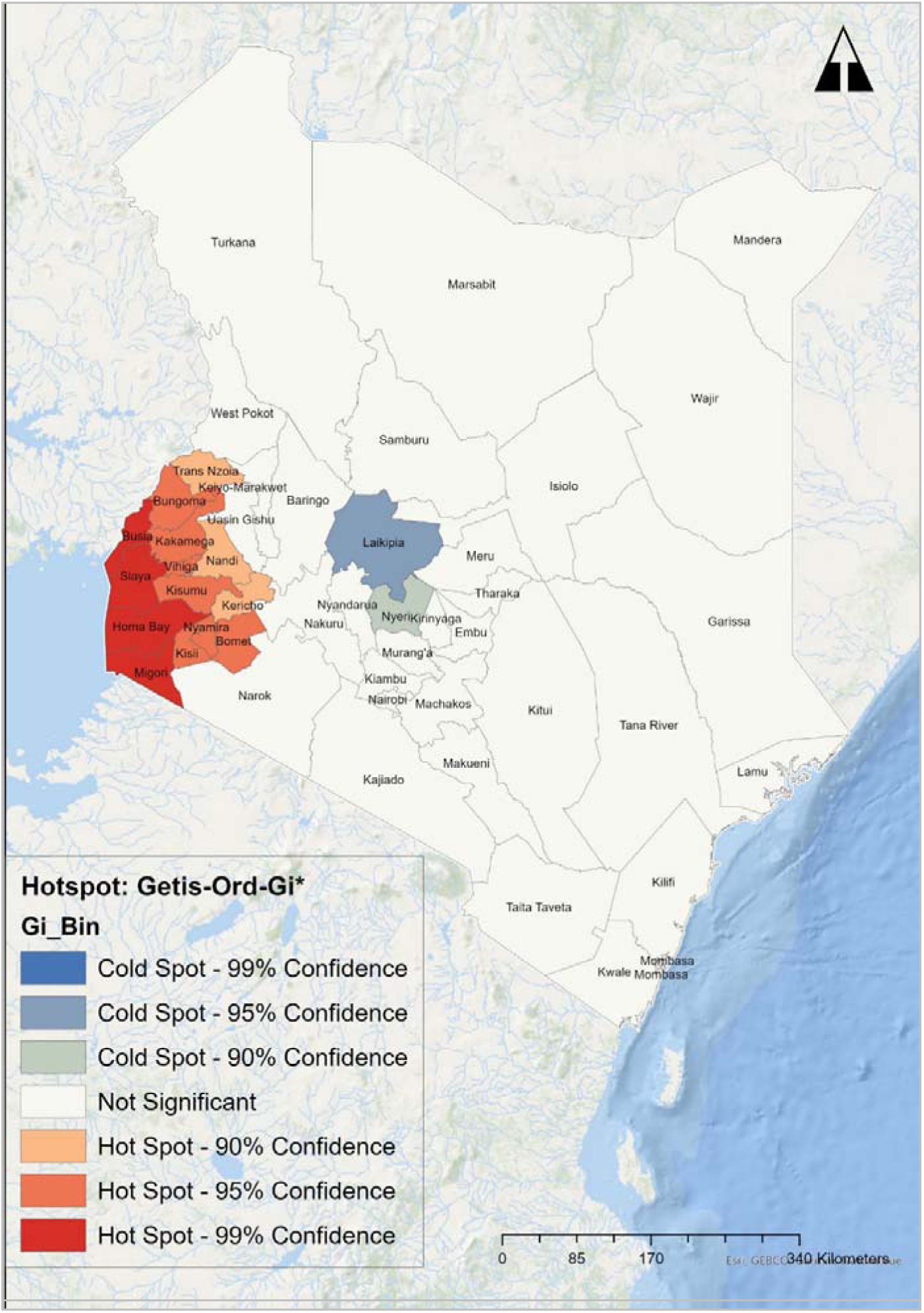
Hotspot Analysis of Predicted Paediatric HIV Positive Cases in Kenya, June 2023.

Spatial autocorrelation analysis confirmed statistically significant clustering of paediatric HIV cases. For reported cases, Moran’s I = 0·22 (z = 3·23, p = 0·001), and for predicted cases, Moran’s I = 0·37 (z = 4·92, p < 0·001). These results indicate that counties with higher paediatric HIV case burdens tend to be geographically proximate, reflecting non-random spatial concentration. The stronger clustering in predicted data suggests that ML estimation sharpened the detection of underlying spatial patterns.

Hotspot analysis using the Getis-Ord Gi* statistic identified significant high-intensity clusters in western Kenya surrounding Lake Victoria, particularly in Homa Bay, Siaya, and Kisumu counties. These hotspots correspond with areas of known high HIV case burden and service demand, validating the analytic framework. Cold-spot areas with low paediatric HIV case numbers were concentrated in central counties such as Laikipia and Nyeri. Together, these spatial findings highlight the need for geographically differentiated responses, prioritizing intensified paediatric case finding and treatment support in high-burden western zones while maintaining preventive gains elsewhere.

## Discussion

This study developed and validated a methodological framework that integrates supervised machine-learning prediction with geostatistical analysis to improve the surveillance of paediatric HIV cases in Kenya. The approach accurately predicted county-level case distribution and, by converting predictions to population-adjusted incidence (cases per 10,000 children aged 0–14 years), enabled a more meaningful comparison of paediatric HIV burden across counties of varying size. Significant geographic clusters were identified through both Moran’s I and Getis-Ord Gi* statistics. These findings demonstrate the potential of predictive analytics to complement national HIV surveillance, enabling Kenya’s HIV programme to strengthen paediatric case finding, target prevention resources, and evaluate regional programme performance.

Consistent with previous research, PMTCT HIV seropositivity was the strongest predictor of paediatric HIV burden, underscoring the central role of maternal testing and treatment in reducing vertical transmission. Strengthening PMTCT coverage and adherence remains essential, particularly through improved linkage of antenatal and postnatal care services, expanded community-based testing for pregnant women, and sustained ART follow-up. The observed association between stunting and paediatric HIV cases likely reflects the cumulative impact of infection, malnutrition, and health system gaps rather than a direct causal pathway. Integrated child-health programmes that address both infection management and nutrition could therefore improve paediatric outcomes. Overlapping patterns between higher Fansidar use and paediatric HIV cases suggest that co-endemicity of malaria and HIV in western Kenya may contribute to shared health-service challenges, reaffirming the need for integrated maternal and child health interventions.

Frequent antenatal care visits were associated with lower paediatric HIV case burdens, emphasizing prenatal care as an effective platform for HIV testing, ART initiation, and treatment adherence support. The negative correlation between male partner HIV testing and paediatric HIV burden highlights the benefit of male engagement in PMTCT and family-centered HIV care. Conversely, sexual violence and multiple sexual partnerships were linked to higher paediatric case detection, aligning with recent *Lancet HIV* findings that reducing partner violence improves maternal and child health outcomes.^1^□ Strengthening community-level prevention of gender-based violence and expanding access to sexual and reproductive health education can indirectly reduce paediatric HIV transmission risks.

Inversely correlated variables such as women’s property ownership and regular postnatal care visits provide evidence of protective socioeconomic and health system factors. These results align with previous studies showing that women’s empowerment and continuous care engagement enhance family health outcomes.¹²,¹³ Strengthening these social determinants of health through microeconomic support, social protection programmes, and gender equity interventions can contribute to sustained reductions in paediatric HIV cases.

The hotspot analysis identified consistent spatial clustering in western Kenya, especially in Homa Bay, Kisumu, and Siaya, which have long been recognized as HIV epicenters. Alignment between model predictions and national surveillance data reinforces the validity of this framework and is consistent with earlier spatial analyses of HIV burden in Kenya.□,^1^□ However, elevated predicted cases in some low-burden counties suggest possible underdiagnosis, testing gaps, or emerging transmission patterns. Incorporating these predictive signals into Kenya’s National Data Warehouse (NDW), case-based surveillance (CBS), and DHIS2 analytics could strengthen epidemic monitoring, allowing sub-national teams to validate predictions with real-time programme data and rapidly identify service gaps.

The quantification of county-level uncertainty through bootstrapped prediction intervals (Supplementary Table 2 and Supplementary Figure 1) strengthens the framework’s utility for operational decision-making by allowing counties to be interpreted not simply by their point prediction but by the full range of plausible values consistent with the model’s observed prediction errors. Empirical coverage of 95·7% closely matched the 95% nominal level, indicating that the bootstrap procedure produced well-calibrated uncertainty bounds across Kenya’s 47 counties. The two counties whose reported case counts fell outside their 95% prediction intervals (Homa Bay and Siaya) both remain historically high-burden paediatric HIV settings in western Kenya, yet exhibited divergent patterns that carry distinct programmatic implications. In Homa Bay, the reported case count exceeding the upper prediction bound is consistent with intensified programmatic case-finding — likely reflecting PEPFAR-supported index testing, enhanced PMTCT follow-up, and community-based paediatric HIV outreach — detecting paediatric HIV cases at a rate beyond what county-level demographic and programmatic indicators alone would predict. In Siaya, by contrast, the reported count falling below the lower prediction bound may indicate gaps in case detection relative to the model-expected burden and warrants targeted investigation of testing coverage and linkage-to-care pathways. This differential interpretation, which emerges directly from the uncertainty analysis rather than from point predictions alone, illustrates how combining penalised predictive modelling with formal uncertainty quantification can move routine surveillance from retrospective case counting toward prospective identification of counties where programmatic intensification is either succeeding or where closer scrutiny may be warranted.

This framework also has broader implications for epidemic control in sub-Saharan Africa. The use of population-adjusted incidence rather than raw case counts represents an important analytical refinement: absolute numbers can overstate burden in densely populated counties while masking disproportionate risk in smaller, underserved ones. Presenting results as incidence per 10,000 paediatric population enables direct inter-county comparison and more equitable resource allocation. When integrated into Kenya’s digital health infrastructure, these predictive models can support adaptive decision-making for epidemic response, programmatic monitoring, and national planning. Similar applications could extend to other infectious-disease programmes seeking to optimise geographic targeting of limited resources.

This study has limitations. The analysis relied on secondary aggregate data, which may include reporting inconsistencies. The modest sample size (n = 47 counties) relative to the 19 candidate predictors constrained statistical power for individual coefficient estimation; penalised regression with L1 and L2 shrinkage mitigates overfitting in this setting, but rankings of individual predictor importance should be interpreted as exploratory rather than definitive. The residual-based bootstrap prediction intervals assume that prediction errors are exchangeable across counties; this assumption may be violated in the presence of spatially structured residuals, in which case the reported intervals could systematically under- or over-cover in specific regions. While the model identifies associations, it cannot infer causality. Geographic aggregation at the county level may mask local heterogeneity, and finer-scale analyses could reveal additional spatial nuances. The population denominators used for incidence calculations were derived from census projections rather than direct enumeration, introducing a potential source of imprecision, particularly for counties with high migration or displacement. Nonetheless, the approach provides a valuable, scalable framework that enhances understanding of paediatric HIV distribution and can inform targeted interventions within existing surveillance systems.

## Conclusion

This study demonstrates the value of combining machine learning prediction with spatial analytics to enhance paediatric HIV surveillance in Kenya. The framework provides a replicable model for data-driven, geographically targeted epidemic control, enabling health authorities to focus on interventions where they are most needed. By integrating predictive outputs into national systems such as DHIS2, NDW, and CBS, Kenya can strengthen paediatric case finding, optimise PMTCT and index-testing strategies, and allocate resources more efficiently. Continued application of predictive analytics and regular model updates will be vital for accelerating progress toward paediatric HIV elimination and for reinforcing health-system responsiveness across sub-Saharan Africa.

## Contributors

AAO conceived and designed the study, developed the methodology, led data management, conducted data analysis, and wrote the initial manuscript draft. BM, FM, EO, and PY provided technical reviews and proofread the manuscript. All authors (AAO, BM, FM, EO, and PY) reviewed the final manuscript and accept responsibility for the decision to submit for publication.

## Data sharing statement

The aggregate-level PEPFAR HIV program data from the Kenya national HIV program and the 2022 Kenya DHS used in this study are publicly accessible. The PEPFAR data can be accessed through the PEPFAR Panorama Spotlight (https://data.pepfar.gov), and the DHS data are available from the DHS Program website (https://dhsprogram.com/). The code used to develop and implement the ML model will be made available by the corresponding author upon reasonable request, subject to agreement on data privacy and appropriate use. Researchers interested in replicating or building upon this study are encouraged to contact the authors for further information on data access and code availability.

## Declaration of interests

In this study, we report no financial or non-financial competing interests. All other authors report no disclosures.

## Acknowledgments

We acknowledge the technical teams from the Kenya Centers for Disease Control and the U.S. Department of State Office of Foreign Assistance Health in Nairobi for their collaboration and support throughout this study. Their contributions and technical insights helped strengthen the quality and relevance of this work and supported efforts to advance understanding of paediatric HIV in Kenya.

## Supplementary Materials

**Supplementary Table 1.** Spatial clustering of paediatric HIV cases in Kenya: Getis–Ord Gi* hotspot analysis. Counties classified as statistically significant hotspots or coldspots of paediatric HIV cases based on Getis–Ord Gi* statistics, October 2022 – June 2023. **Notes:** Hotspots are counties with statistically significant clustering of elevated paediatric HIV case counts among neighbouring counties, identified using the Getis–Ord Gi* statistic. Coldspots are counties with statistically significant clustering of low paediatric HIV case counts. Confidence levels: 99% (p<0.01), 95% (p<0.05), and 90% (p<0.10). Positive z-scores indicate hotspots; negative z-scores indicate coldspots. Analysis was conducted on 47 Kenyan counties using queen contiguity spatial weights in ArcGIS Pro v3.1.

| Rank | County | $G_i^*$ z-score | p-value | Classification | Confidence |
| --- | --- | --- | --- | --- | --- |
| 1 | Migori | +3.40 | 0.0006 | Hotspot | 99% |
| 2 | Siaya | +3.01 | 0.0020 | Hotspot | 99% |
| 3 | Homa Bay | +2.98 | 0.0020 | Hotspot | 99% |
| 4 | Busia | +2.75 | 0.0050 | Hotspot | 99% |
| 5 | Kisii | +2.55 | 0.010 | Hotspot | 95% |
| 6 | Bungoma | +2.51 | 0.012 | Hotspot | 95% |
| 7 | Kakamega | +2.39 | 0.016 | Hotspot | 95% |
| 8 | Kisumu | +2.23 | 0.025 | Hotspot | 95% |
| 9 | Bomet | +2.05 | 0.039 | Hotspot | 95% |
| 10 | Laikipia | -2.03 | 0.041 | Coldspot | 95% |
| 11 | Vihiga | +2.16 | 0.045 | Hotspot | 95% |
| 12 | Nandi | +1.91 | 0.055 | Hotspot | 90% |
| 13 | Kericho | +1.82 | 0.067 | Hotspot | 90% |
| 14 | Nyeri | -1.76 | 0.078 | Coldspot | 90% |
| 15 | Trans Nzoia | +1.76 | 0.077 | Hotspot | 90% |

The analysis identified 13 counties in western Kenya (Migori, Siaya, Homa Bay, Busia, Kisii, Bungoma, Kakamega, Kisumu, Bomet, Vihiga, Nandi, Kericho, and Trans Nzoia) as statistically significant hotspots of elevated paediatric HIV burden. Laikipia and Nyeri counties were identified as statistically significant coldspots, with lower paediatric HIV case counts than expected under a null hypothesis of spatial randomness. These findings support geographically targeted intensification of paediatric case-finding and treatment services in western Kenya, while informing differentiated service delivery across the remaining counties.

**Supplementary Figure 1.**
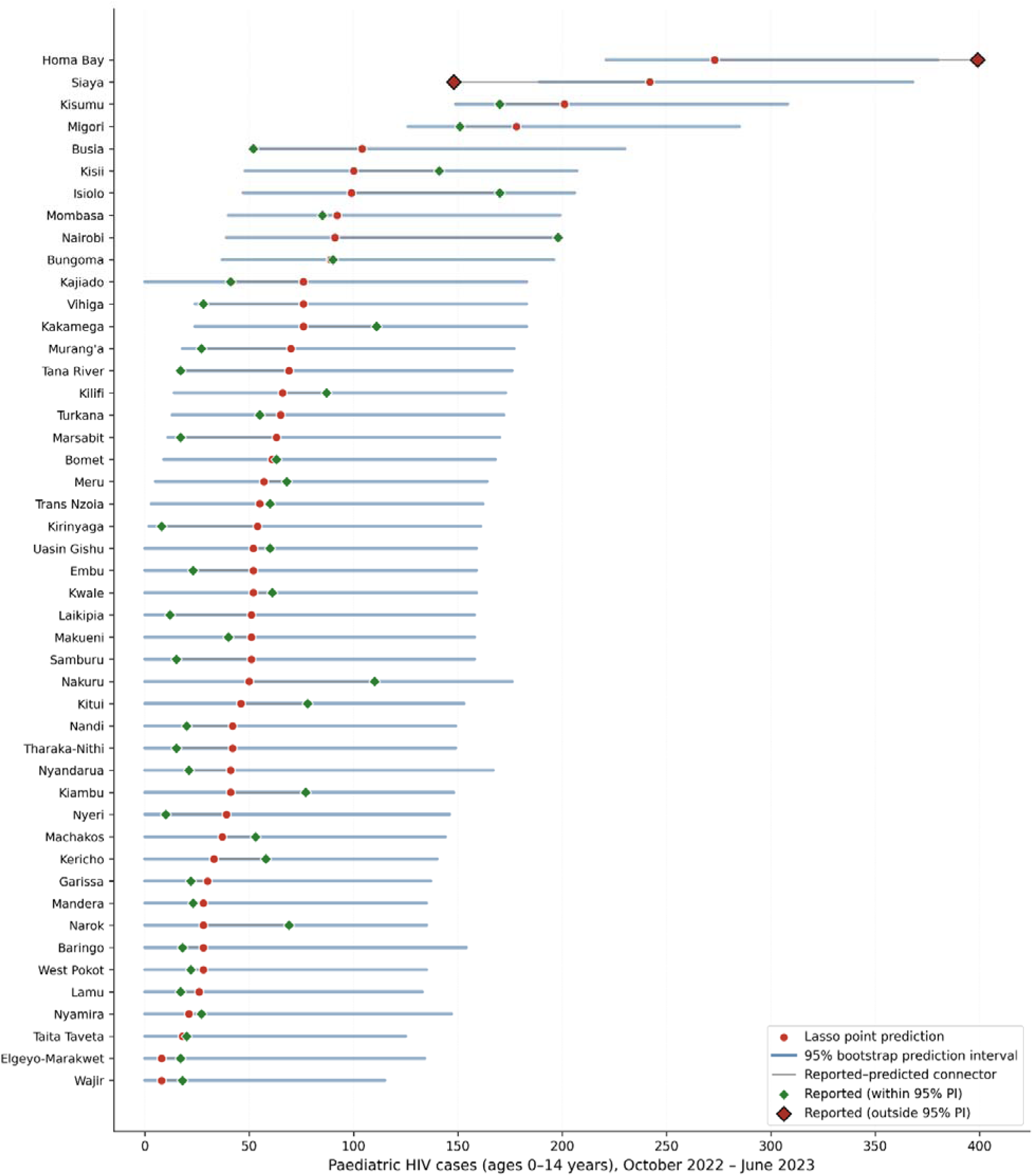
County-level paediatric HIV cases across 47 Kenyan counties: reported values, Lasso predictions, and bootstrapped 95% prediction intervals, October 2022 – June 2023.

Consolidated forest plot displaying, for each of Kenya’s 47 counties, four elements: (i) the Lasso model’s point prediction for paediatric HIV case count (red dot); (ii) the 95% bootstrap prediction interval around that point prediction (blue horizontal band); (iii) the reported case count from the national HIV testing programme (green diamond if within the prediction interval, coral diamond with black outline if outside); and (iv) a thin grey connector line between the predicted and reported values, allowing direct visual assessment of the reported–predicted discrepancy at each county. Counties are ordered from highest to lowest predicted case count. National totals: 3,092 reported versus 3,160 predicted cases (difference = 2.2%). Across the 47 counties, reported case counts fell within the 95% prediction interval for 45 counties (empirical coverage 95.7%, closely matching the nominal 95% level), with a median prediction interval width of 159 cases. Two counties are flagged as outliers: Homa Bay, whose reported case count (399) exceeds the upper bound of the prediction interval (380), and Siaya, whose reported count (148) falls below the lower bound (189). Both are historically recognised as among Kenya’s highest-burden paediatric HIV settings; the directions of their respective outlying values suggest distinct programmatic signals, discussed in the main text.

**Supplementary Table 2.** Lasso-predicted county-level paediatric HIV cases with bootstrapped 95% prediction intervals, Kenya (October 2022 – June 2023) County-level point predictions and 95% prediction intervals derived from 2000 residual-based bootstrap iterations of the tuned Lasso regression model; residuals resampled with replacement and added to fitted values to propagate outcome-level uncertainty. Counties ordered by predicted case count.

| Rank | County | Reported | Predicted | PI Lower | PI Upper | PI Width | Incidence (/10,000) | Within 95% PI |
| --- | --- | --- | --- | --- | --- | --- | --- | --- |
| 1 | Homa Bay | 399 | 273 | 221 | 380 | 159 | 5.25 | Outside |
| 2 | Siaya | 148 | 242 | 189 | 368 | 179 | 5.71 | Outside |
| 3 | Kisumu | 170 | 201 | 149 | 308 | 159 | 4.28 | Within |
| 4 | Migori | 151 | 178 | 126 | 285 | 159 | 3.32 | Within |
| 5 | Busia | 52 | 104 | 51 | 230 | 179 | 2.65 | Within |
| 6 | Kisii | 141 | 100 | 48 | 207 | 159 | 1.96 | Within |
| 7 | Isiolo | 170 | 99 | 47 | 206 | 159 | 6.54 | Within |
| 8 | Mombasa | 85 | 92 | 40 | 199 | 159 | 2.19 | Within |
| 9 | Nairobi | 198 | 91 | 39 | 198 | 159 | 0.62 | Within |
| 10 | Bungoma | 90 | 89 | 37 | 196 | 159 | 1.16 | Within |
| 11 | Vihiga | 28 | 76 | 24 | 183 | 159 | 3.34 | Within |
| 12 | Kakamega | 111 | 76 | 24 | 183 | 159 | 0.96 | Within |
| 13 | Kajiado | 41 | 76 | 0 | 183 | 183 | 1.51 | Within |
| 14 | Murang'a | 27 | 70 | 18 | 177 | 159 | 2.03 | Within |
| 15 | Tana River | 17 | 69 | 17 | 176 | 159 | 4.17 | Within |
| 16 | Kilifi | 87 | 66 | 14 | 173 | 159 | 1.00 | Within |
| 17 | Turkana | 55 | 65 | 13 | 172 | 159 | 1.48 | Within |
| 18 | Marsabit | 17 | 63 | 11 | 170 | 159 | 2.52 | Within |
| 19 | Bomet | 63 | 61 | 9 | 168 | 159 | 1.61 | Within |
| 20 | Meru | 68 | 57 | 5 | 164 | 159 | 1.05 | Within |
| 21 | Trans Nzoia | 60 | 55 | 3 | 162 | 159 | 1.26 | Within |
| 22 | Kirinyaga | 8 | 54 | 2 | 161 | 159 | 3.05 | Within |
| 23 | Embu | 23 | 52 | 0 | 159 | 159 | 2.68 | Within |
| 24 | Uasin Gishu | 60 | 52 | 0 | 159 | 159 | 1.15 | Within |
| 25 | Kwale | 61 | 52 | 0 | 159 | 159 | 1.24 | Within |
| 26 | Samburu | 15 | 51 | 0 | 158 | 158 | 3.03 | Within |
| 27 | Laikipia | 12 | 51 | 0 | 158 | 158 | 2.43 | Within |
| 28 | Makueni | 40 | 51 | 0 | 158 | 158 | 1.48 | Within |
| 29 | Nakuru | 110 | 50 | 0 | 176 | 176 | 0.57 | Within |
| 30 | Kitui | 78 | 46 | 0 | 153 | 153 | 1.03 | Within |
| 31 | Tharaka-Nithi | 15 | 42 | 0 | 149 | 149 | 3.23 | Within |
| 32 | Nandi | 20 | 42 | 0 | 149 | 149 | 1.18 | Within |
| 33 | Kiambu | 77 | 41 | 0 | 148 | 148 | 0.48 | Within |
| 34 | Nyandarua | 21 | 41 | 0 | 167 | 167 | 1.83 | Within |
| 35 | Nyeri | 10 | 39 | 0 | 146 | 146 | 1.75 | Within |
| 36 | Machakos | 53 | 37 | 0 | 144 | 144 | 0.74 | Within |
| 37 | Kericho | 58 | 33 | 0 | 140 | 140 | 0.90 | Within |
| 38 | Garissa | 22 | 30 | 0 | 137 | 137 | 0.60 | Within |
| 39 | Baringo | 18 | 28 | 0 | 154 | 154 | 0.91 | Within |
| 40 | West Pokot | 22 | 28 | 0 | 135 | 135 | 0.83 | Within |
| 41 | Mandera | 23 | 28 | 0 | 135 | 135 | 0.56 | Within |
| 42 | Narok | 69 | 28 | 0 | 135 | 135 | 0.45 | Within |
| 43 | Lamu | 17 | 26 | 0 | 133 | 133 | 4.22 | Within |
| 44 | Nyamira | 27 | 21 | 0 | 147 | 147 | 0.92 | Within |
| 45 | Taita Taveta | 20 | 18 | 0 | 125 | 125 | 1.51 | Within |
| 46 | Elgeyo-Marakwet | 17 | 8 | 0 | 134 | 134 | 0.40 | Within |
| 47 | Wajir | 18 | 8 | 0 | 115 | 115 | 0.19 | Within |

**Notes:** Prediction intervals were generated by 2000 residual-based bootstrap iterations (seed = 20260420), in which the empirical residual distribution (reported minus predicted case count, across all 47 counties) was resampled with replacement and added to each county's point prediction. The procedure preserves the point predictions from the tuned Lasso model. Lower bounds truncated at 0 because case counts cannot be negative. Of 47 counties, reported case counts fell within the 95% prediction interval for 45 counties (95.7% empirical coverage vs 95% nominal). Two counties (shown in coral) had reported case counts outside the 95% PI: Homa Bay (reported 399 vs upper bound 380) and Siaya (reported 148 vs lower bound 189). Median PI width = 159 cases. "Within" indicates the reported value fell inside the 95% PI; "Outside" indicates it fell beyond either bound. PI = prediction interval.

## Notes

### Competing Interest Statement

The authors have declared no competing interest.

### Summary of Updates

The following coauthor and affiliated institutional information have been removed from this version of the manuscript following institutional publication clearance requirements:Gonza Omoro, Jonah Maswai, John Owuoth, Duncan Kirui, and Lydia Odero.

## References

1. UNAIDS. Global HIV & AIDS statistics — fact sheet. Geneva: Joint United Nations Programme on HIV/AIDS, 2021. Available from: https://www.unaids.org/en/resources/fact-sheet

2. UNAIDS. 90-90-90: an ambitious treatment target to help end the AIDS epidemic. Geneva: Joint United Nations Programme on HIV/AIDS, 2014. Available from: https://www.unaids.org/sites/default/files/media_asset/90-90-90_en.pdf

3. Mutanga JN, Ronan A, Powis KM. Achieving equity for children and adolescents with perinatal HIV exposure: an urgent need for a paradigm shift. J Int AIDS Soc 2023; 26(Suppl 4): e26171. doi:10.1002/jia2.26171

4. UNAIDS. Progress in preventing new HIV infections among children. Geneva: Joint United Nations Programme on HIV/AIDS, 2023. Available from: https://www.unaids.org/en/resources/presscentre/featurestories/2023/june/20230616_progress-report

5. Mugo C, Wang J, Begnel ER, et al. Home- and clinic-based pediatric HIV index case testing in Kenya: uptake, HIV prevalence, linkage to care, and missed opportunities. J Acquir Immune Defic Syndr 2020; 85(5): 535–542. doi:10.1097/QAI.0000000000002500

6. Dwyer-Lindgren L, Cork MA, Sligar A, et al. Mapping HIV prevalence in sub-Saharan Africa between 2000 and 2017. Nature 2019; 570(7760): 189–193. doi:10.1038/s41586-019-1200-9

7. Waruru A, Achia TNO, Muttai H, et al. Spatial-temporal pattern of HIV diagnoses in Kenya: implications for intervention and prevention. BMC Public Health 2021; 21: 1171. doi:10.1186/s12889-021-11130-y

8. World Bank. Population density (people per sq. km of land area) - Kenya. World Development Indicators. Washington, DC: World Bank; 2023. Available from: https://data.worldbank.org/indicator/EN.POP.DNST?locations=KE

9. United Nations Children’s Fund (UNICEF). The state of the world’s children 2023: children in a digital world. New York: UNICEF, 2023. Available from: https://www.unicef.org/reports/state-of-worlds-children-2023

10. National AIDS Control Council (NACC). Kenya HIV estimates report 2018. Nairobi: Ministry of Health, 2018. Available from: https://nacc.or.ke/wp-content/uploads/2018/11/HIV-estimates-report-Kenya-20182.pdf

11. World Health Organization. WHO encourages countries to adapt HIV testing strategies in response to changing epidemiology. Geneva: World Health Organization, 2019. Available from: https://www.who.int/publications/i/item/WHO-CDS-HIV-19.34

12. Dunbar MS, Maternowska MC, Kang MS, Laver SM, Mudekunye-Mahaka I, Padian NS. Findings from SHAZ! A feasibility study of a microcredit and life skills HIV prevention intervention to reduce risk among adolescent female orphans in Zimbabwe. J Prev Interv Community 2010; 38(2): 147–161. doi:10.1080/10852351003640849

13. Ditekemena J, Koole O, Engmann C, et al. Determinants of male involvement in maternal and child health services in sub-Saharan Africa: a review. Reprod Health 2012; 9(1): 32. doi:10.1186/1742-4755-9-32

14. Dunkle KL, Decker MR. Addressing partner violence to end infant HIV infection. Lancet HIV 2024; 11(5): e261–e262. doi:10.1016/S2352-3018(24)00180-2

15. Boerma T, Williams BG, Zaidi I. Monitoring the HIV epidemic in sub-Saharan Africa: data gaps and opportunities for more effective use of data. J Glob Health 2016; 6(2): 020201. doi:10.7189/jogh.06.020201

16. Mumo J, Ochieng C, Mwangi J, et al. Leveraging electronic medical records for HIV testing, care, and treatment in Kenya. BMC Med Inform Decis Mak 2023; 23: 265. doi:10.1186/s12911-023-02265-6

